# Deep learning optimisation for cardiology: Neural Architecture Search-driven arrhythmia classification with electrocardiograms

**DOI:** 10.64898/2026.05.28.26354348

**Authors:** Erik Vanegas Müller, Arese Joe-Oshodi, Abhirup Banerjee, Mauricio Villarroel

**Affiliations:** The Podium Institute for Sports Medicine and Technology, Department of Engineering Science, University of Oxford, Oxford, United Kingdom; Institute of Biomedical Engineering, Department of Engineering Science, University of Oxford, Oxford, United Kingdom; Division of Cardiovascular Medicine, Radcliffe Department of Medicine, University of Oxford, Oxford, United Kingdom

**Keywords:** Neural Architecture Search, Deep Learning, Electrocardiogram, Arrhythmia Classification, Self-attention

## Abstract

Cardiovascular disease is the leading cause of death worldwide. Sudden cardiac death (SCD) accounts for roughly 50% of all cardiac deaths. The electrocardiogram (ECG) is widely used for early diagnosis of cardiac disease. However, the complexity of accurate interpretation limits the ECG’s efficacy. Modern deep learning methods have been applied to assist clinicians in diagnosis. We applied Neural Architecture Search (NAS), an automated machine learning technique, to identify optimal deep learning architectures for classifying cardiac arrhythmias from ECGs.

We applied the Differentiable Architecture Search strategy to an AutoFormer search space to identify optimal self-attention architectures for arrhythmia classification. We trained, validated, and tested the resulting model on the PhysioNet Challenge 2021 dataset (n = 88,253), comprising ECGs across three continents. We performed a hyperparameter optimisation on the NAS output, exploring input patch size, class weighting, and loss function. We evaluated performance using the PhysioNet Challenge metric and the area under the receiver operating characteristic curve (AUROC).

The NAS converged towards minimal architectural configurations (embedding dimension: 384, depth: 4, self-attention heads: 4, MLP ratio: 1) with a validation challenge metric of 0.66 (PhysioNet Challenge 21 Winner: 0.63). The NAS-created network achieved an AUROC of 0.97 and a challenge metric of 0.71 during testing. Normal Sinus Rhythm and Sinus Tachycardia achieved AUROCs of 0.99. Low-QRS Voltage and T-wave abnormality were the worst-performing arrhythmias, with AUROCs of 0.89 and 0.90, respectively.

We interpret that architectural simplicity drives performance in arrhythmia classification. Because SCD is unexpected, prevention strategies in free-living environments require lightweight computational resources suitable for wearable devices. Class imbalance fundamentally limits classification performance for rare arrhythmias such as Low-QRS Voltage and T-wave inversion, irrespective of hyperparameter choices. However, the self-attention mechanism can autonomously abstract clinical representations, simplifying clinical deployment by eliminating the need for an explicit feature-extraction pipeline.

**Author summary:** Heart diseases are the leading cause of death worldwide, of which around 50% happen suddenly. Analysis of the heart’s electrical signal can help detect heart disease early. However, it is a difficult and time-consuming process, even for experienced clinicians. Artificial intelligence (AI) can assist clinicians in detecting heart disease. We explore a specialised form of artificial intelligence that automatically finds its own optimal structure. To do this, we used a dataset from the general population containing 88,253 heart’s electrical activity records, commonly called electrocardiograms.

We found that simpler AI architectures perform better at identifying heart disease. This finding is contrary to the very prevalent thought “the bigger the better”, making this technology ideal for wearables, considering that almost 50% of the heart disease deaths are sudden. This finding is significant since simpler AI architectures require much less computing power and battery life, with the potential to make heart disease monitoring more accessible and equitable for everyone, especially in low-resource settings. Furthermore, we reduce human bias and the trial-and-error that often follows when creating optimised structures.

## Introduction

Cardiovascular disease is the leading cause of death worldwide, accounting for approximately 20.5 million deaths globally annually and expected to continue rising to 35.6 million in 2050 . Sudden cardiac death (SCD), defined as a death within one hour of the onset of symptoms (presumably of cardiac origin), accounts for roughly 50% of all worldwide cardiac deaths . The largest absolute number of SCD cases occurs in the general population, who are unaware of their risk . Abnormalities found on an Electrocardiogram (ECG) are predictive of early and late SCD events in middle-aged subjects .

The clinical interpretation and diagnosis of cardiac abnormalities on the ECG are both time-consuming and require highly trained clinicians who practice ECG reading in a consistent and systematic manner . These challenges are further compounded by the existence of over 100 different types of cardiac abnormalities, with varying degrees of difficulty in identification . Multiple diagnostic algorithms have been developed to aid clinicians in interpreting ECGs. However, these algorithms can be inaccurate in terms of sensitivity and specificity, vary widely in their evaluation of ECG recordings, and lack diagnostic robustness . Diagnostic algorithms, therefore, can introduce additional uncertainty into a clinician’s diagnosis, delay treatment, and hamper clinicians’ assessment .

Applications of deep learning models to ECG interpretation have shown significant improvements in accuracy and diagnostic robustness compared to traditional algorithms . The European Heart Rhythm Association of the European Society of Cardiology (ESC), the Heart Rhythm Society (HRS), and the ESC Working Group on E-Cardiology identified AI in sudden cardiac death as a key research area in AI applied to Clinical Electrophysiology (the other being AI in atrial fibrillation management and AI in the electrophysiology lab) .

Barker et al. used a residual neural network to predict from 24h ambulatory ECGs whether patients would experience ventricular arrhythmia. Of 270 analysed patients, 159 had a ventricular arrhythmia outcome. The deep learning model reached an accuracy of 76%. Holmstrom et al. trained a deep learning model on 1,827 pre-cardiac arrest 12-lead ECGs from 1,796 SCD cases, along with a control group of 1,342 ECGs from 1,325 individuals (at least 50% of whom had established coronary artery disease). The model achieved an Area Under the Receiver Operating Characteristic (AUROC) of 0.889 on an internal test set and 0.820 on an external validation test set with 714 SCD cases.

AI tools could therefore enable personalised SCD risk prediction, for which the choice of deep learning architecture is decisive. The architecture’s design depends on the researchers’ prior knowledge and experience, and is heavily problem-dependent . Therefore, the design choices are not necessarily interchangeable between different fields of arrhythmia classification applications. The field of neural architecture search (NAS) aims to automate the construction of deep learning architectures, thereby reducing the potential biases introduced by traditional manual architecture design and hyperparameter fine-tuning processes .

Azarmehr et al. designed a neural architecture search for echocardiography view classifiers, reducing trainable parameters by up to 99.9% while maintaining comparable classification performance. However, most NAS approaches have been developed and validated on image classification tasks, with limited exploration of physiological time-series data such as ECGs.

We apply NAS to identify an optimal deep learning architecture for arrhythmia classification. We use the PhysioNet Challenge 2021 dataset and represent the 12-lead ECGs as 12-channel inputs. We compare the performance of a NAS-derived architecture against a conventional manually-designed baseline model (PhysioNet Challenge 2020 winner) across different arrhythmias. We hypothesise that automated architecture search identifies architectures with superior or comparable classification performance, replacing costly manual trial-and-error with a single optimised search.

## Methods

### Dataset

We use the PhysioNet 2021 dataset for the NAS-driven arrhythmia classification. The PhysioNet Challenge 2021 ECG recordings originate from seven different institutions on three continents (America, Asia, Europe). The publicly available dataset includes 88,253 ECGs with 133 diagnoses or classes. Of these 133 classes, 30 (see fig. 1 for a selection of four arrhythmias) are used to evaluate the submitted algorithms for the PhysioNet challenge 2021. The choice of those 30 arrhythmias is justified by their clinical frequency and relevance, as well as their likelihood of being recognisable from ECG recordings . The patient’s mean age in the dataset is 59.7 ± 21.7 years. The sex distribution is 43.5% female and 56.5% male.

**Figure 1.**
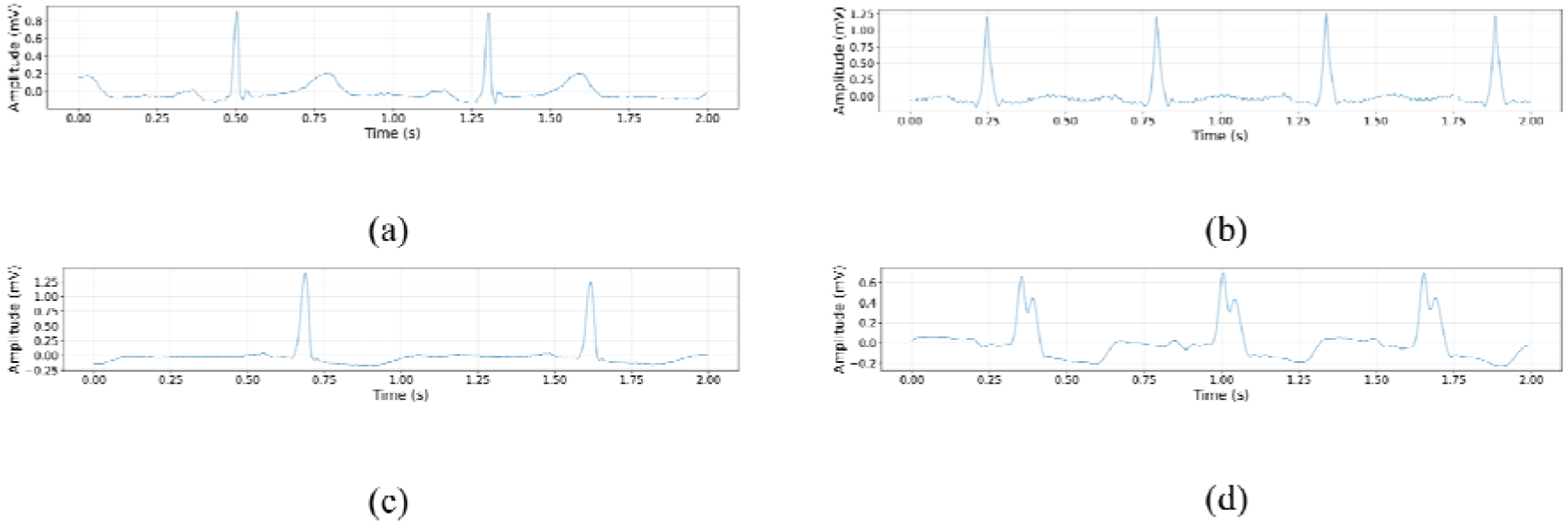
Representative 2-second ECG segments for four arrhythmia classes in the PhysioNet Challenge 21 dataset. (a) Record E04837 lead II showing normal sinus rhythm. The patient is in its 50s and female. (b) Record JS00561 lead II demonstrating sinus tachycardia. The patient is in its 20s and male. (c) Record E10121 lead V5 exhibiting T-wave inversion. The patient is in its 60s and male. (d) Record Q2170 lead V2 displaying right bundle branch block morphology. The patient is in its 80s and male.

### Deep learning architecture

We base our deep learning architecture on Natarajan et al. . The network architecture comprises a “clinical” and a “raw signal” features transformer for multi-label ECG classification. The “clinical” features comprise the 20 most critical cardiac biomarkers—such as heart rate or RR interval—selected by an initial random forest model from more than 300 ECG features. The “raw signal” features are directly extracted from the ECG waveform. The “raw signal” learned features are passed to a transformer head. After the transformer head, we combine the “clinical” and “raw signal” features into the final fully connected layer.

We chose the challenge metric from the PhysioNet Challenge as the primary performance metric. The challenge metric converts the multi-label classification problem (i.e., multiple labels per ECG) into a multi-class evaluation system (i.e., one primary condition per ECG). The challenge metric gives full credit for correct classifications and partial credit for incorrect classifications, with closely related conditions receiving more credit than those that are entirely unrelated (15). The area under the receiver operating characteristic curve (AUROC) is our secondary performance metric.

#### Preprocessing

We filter the ECG signal with a bandpass Finite Impulse Response filter (bandwidth 3Hz to 45Hz, order 150) for noise reduction. Then, we apply min-max normalisation to constrain the lead recordings to the range −1 to 1. The min-max normalisation is distribution-agnostic and preserves the relative importance of values in the data. We resample all ECG recordings to 500Hz and set the sequence length to 15s. Sequence lengths shorter than 15s were zero-padded, whilst a 15s window was taken out of longer signals.

We divide the ECG signal into uniform patches of 0.05s (25 points at 500Hz) to ensure compatibility with the vision transformer architecture. We chose the patch size to be smaller than the QRS complex duration (≤ 0.10s in adults), the fastest-changing component of the cardiac waveform, to ensure we capture rapid ventricular transitions during time-series ECG-to-transformer-required patch-format conversion .

We split the publicly available dataset into 80% for NAS training, 10% for validation, and 10% for testing and employ iterative stratification to ensure equal distribution of cardiac abnormalities across folds . The 80% NAS training dataset is further split into a 50% NAS training set and a 50% NAS validation set following the methodology for DARTS search proposed by Liu, Simonyan, and Yang .

#### Search space

A NAS consists of a search space, a search strategy, and a performance estimation metric . Four search spaces exist: Macro, cell, hierarchical, and chain-structured. Macro search spaces explore a large number of architectures . Cell-based search spaces exhibit limited variance in predictive capability, regardless of their design . Hierarchical search spaces pose significant technical implementation challenges . A chain-structured search space provides a suitable compromise between computational feasibility and architectural variation.

We utilise the “AutoFormer” chain-structured search space from the Neural Network Intelligence (NNI). AutoFormer addresses self-attention blocks and convolutional layers and is therefore well-suited to our network’s architecture . The AutoFormer search space can be subdivided into macro and micro architecture search spaces, where parameters involving macro architecture design choices are more memory-intensive than those involving micro architecture design choices (see fig. 2).

**Figure 2.**
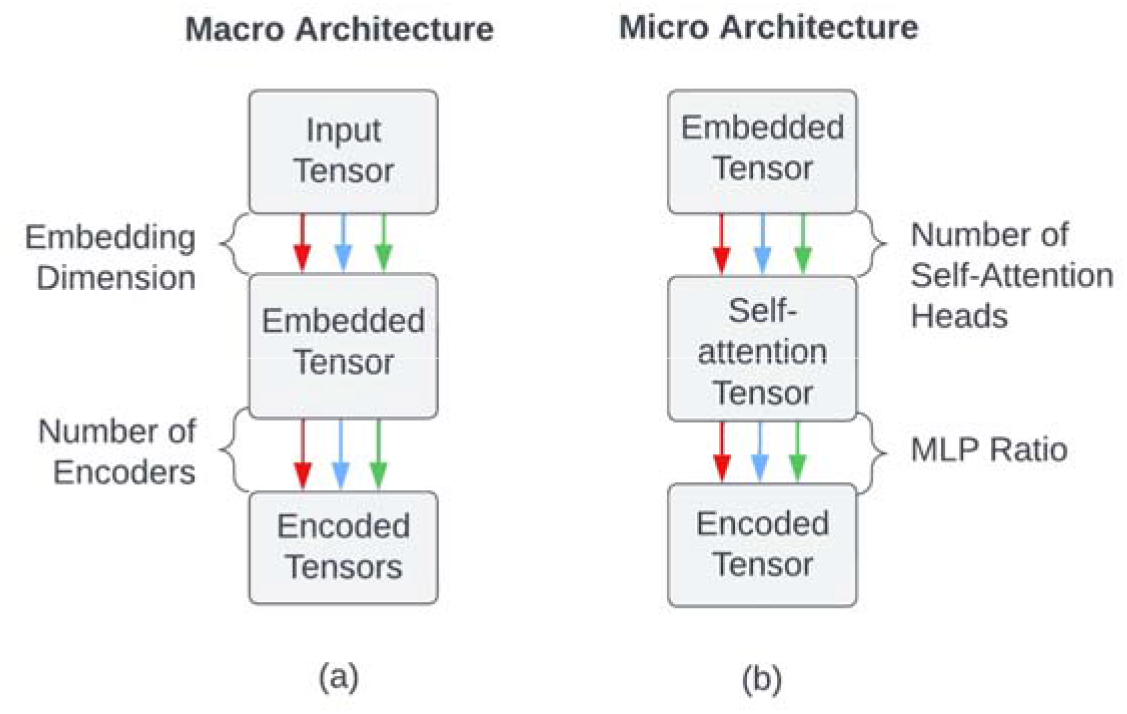
Relaxed directed acyclic graph of the (a) macro architecture search space and (b) micro search space. Grey cells indicate nodes, and the lines between cells represent edges. The different edge colours correspond to the available design options. Annotations exist between cells to indicate the overarching design choice encapsulated by the edge.

We construct a search space comprising 85.9 million parameters. For the Macro architecture parameters, we searched the embedding dimension (384, 768, 1,024) of the input vector and the network’s depth (4, 6, 8), i.e., number of encoders. The number of self-attention heads (4, 6, 8) in an encoder layer, the associated Query, Key, and Value (Q-K-V) dimensions (fixed), and the Multi-Layer Perception (MLP) ratio (1, 2, 4) within an encoder are the microarchitecture search parameters. Deeper vision transformers could have significantly increased the risk of overfitting, given that the PhysioNet 21 dataset is 1.3, 14, and 303 million times smaller than the standard training set sizes for vision-based transformers such as ImageNet-21k, ImageNet, and JFT, respectively .

The AutoFormer search space was developed for image classification and carries no inductive bias towards any specific dataset . We therefore rearrange the one-dimensional ECG into a 12-lead image (i.e., a two-dimensional image). The vision transformer separates input images into patches and embeds them as vectors using either convolutional or linear operations. We add the positional and class encodings to the embedded vectors before passing them into a conventional transformer encoder block. The output from the encoder blocks is passed to a classification block

#### Search strategy

Black-box algorithms offer simpler implementations without compromising search robustness. At the same time, these algorithms are computationally demanding . A one-shot strategy provides a suitable compromise between computational feasibility and search robustness.

We utilise the Differentiable ARchitecTure Search (DARTS) to explore an AutoFormer search space, as it was the most computationally feasible option available in the NNI package. DARTS transforms the discrete decision search space into a continuous, differentiable one through a process known as “relaxation” (see Appendix for more information).

#### Performance estimation

We use the standard binary cross-entropy (BCE) loss function as the primary performance metric for the NAS search, along with an additional elementary macro-accuracy measure.

### Hyperparameter optimisation

We perform hyperparameter optimisation on the NAS output using the PhysioNet 2020 dataset (a subset of the PhysioNet 2021 dataset). We remove any overlapping ECGs between the PhysioNet 2020 and the validation or test folds of the 2021 dataset. We split the PhysioNet 2020 dataset for hyperparameter optimisation into an 80% training, 10% validation, and 10% test folds using iterative stratification (18). The hyperparameters we optimise are the input patch sizes (0.05, 0.1, and 0.2) and the loss function type (class weights, focal loss, and asymmetric loss (ASL); see the Appendix for more information).

We also investigate the inclusion of three sets of clinical features as inputs to the deep model. The first set utilises the output of a random forest model that identified the 20 most important morphological, clinical, and statistical features present in a lead II ECG recording . The second set contains demographic data regarding the patient’s age and gender. The third set is a combination of the first two sets.

## Results

The winner of the PhysioNet Challenge 21 achieved a validation challenge score of 0.63 . Our deep learning architecture achieved an AUROC of 0.95 and a higher challenge metric of 0.66 during validation. During testing, the results were 0.97 for AUROC and 0.71 for the challenge metric. The best scores for individual arrhythmias were Normal Sinus Rhythm, Sinus Tachycardia, and Right Bundle Branch Block, with AUROCs of 0.99. Low-QRS voltage, T-wave abnormality, and T-wave inversion were the worst-performing arrhythmias, with AUROCs of 0.89, 0.90, and 0.92 respectively.

Figure 3 shows the NAS using the DARTS search strategy on the AutoFormer search space over 50 epochs. Each line represents an architecture evaluated during the search, with colour indicating validation accuracy (blue for higher accuracy, red for lower). The visualisation shows architectural parameters (embedding dimension, network depth, and block-specific choices for self-attention heads and MLP ratios) alongside validation loss and accuracy. We display only the first 4 encoder blocks since the search rarely selected deeper networks. We excluded parameter values that were never chosen during the search (e.g., E4 with 6 or 4 heads) from the visualisation. The optimal architecture uses an embedding dimension of 384, a depth of 4 encoder blocks, 4 self-attention heads, and an MLP ratio of 1.

**Figure 3.**
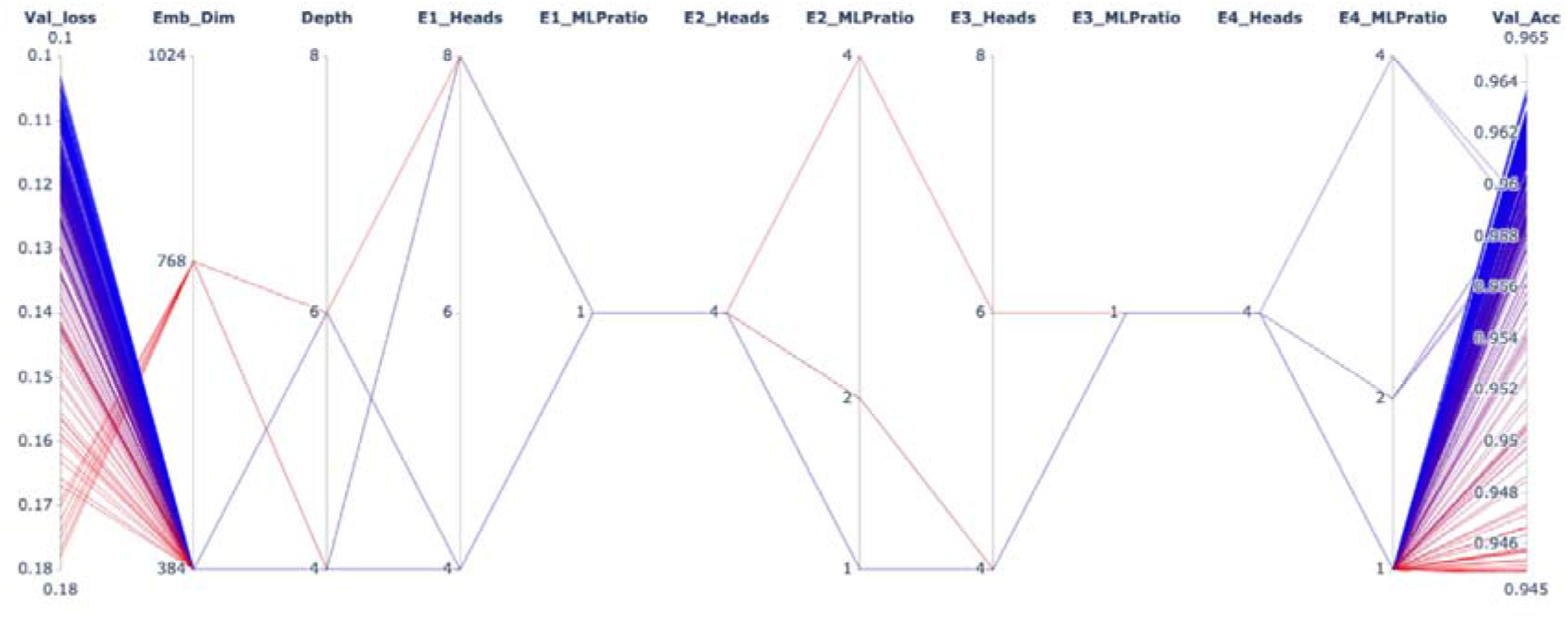
Parallel coordinates visualisation of NAS using DARTS on the AutoFormer search space over 50 epochs. Each line represents an architecture evaluated during the search, with colour indicating validation accuracy (blue for higher, red for lower). The visualisation shows architectural parameters (embedding dimension, network depth, and encoder block configurations including self-attention heads and MLP ratios) alongside validation loss and accuracy. Only the first four encoder blocks (E1-E4) are displayed, as deeper networks were rarely selected. Parameter values never chosen during the search were excluded from the visualisation.

Figure 4 shows the hyperparameter optimisation results using parallel coordinates plots. Each line represents a hyperparameter configuration, with colour indicating the challenge metric score (blue for higher scores, red for lower). The plots display hyperparameter values (patch size, clinical features, loss function, weighted classes, and weight initialisation) alongside validation performance (challenge metric and AUROC). The optimal configuration used a patch size of 25, no clinical features, BCE loss without class weighting, and Xavier initialisation, achieving an AUROC of 0.91 and challenge metric of 0.5.

**Figure 4.**
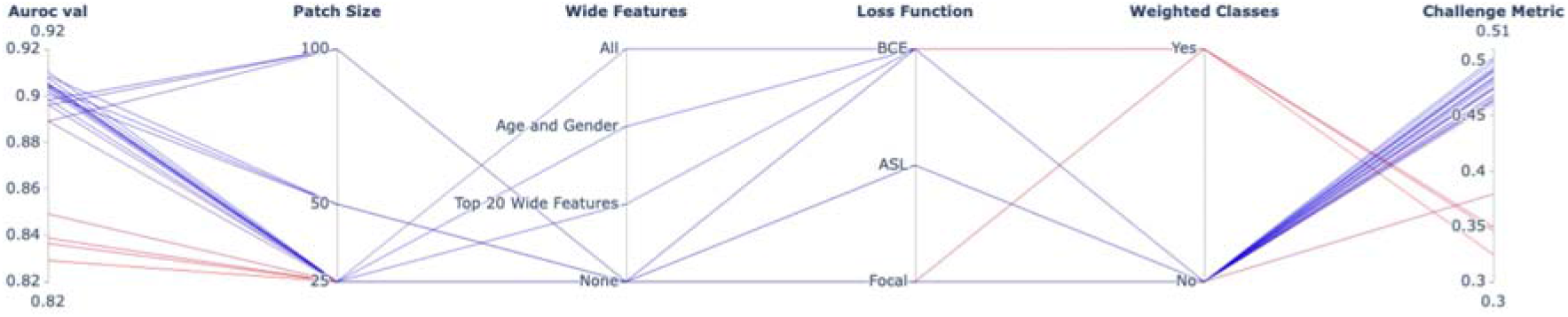
Parallel coordinates visualisation of hyperparameter optimisation results. Each line represents a hyperparameter configuration, with colour indicating challenge metric score (blue for higher, red for lower). The visualisation shows the hyperparameter values for patch size, clinical features (here called wide features as in), loss function, and weighted classes alongside the validation performance metrics AUROC and the challenge metric.

## Discussion

### Neural Architecture Search

We apply NAS to design a deep learning architecture for arrhythmia classification. Early NAS iterations prefer larger architectures, as indicated by an embedding dimension of 768 and depth of 6. However, as the search progresses, both macro and micro architecture parameters converge towards minimal configurations: an embedding dimension of 384, a depth of 4 blocks, 4 self-attention heads, and an MLP ratio of 1. Since DARTS has no preference for minimising architectural parameters, the convergence towards minimal configurations indicates these perform better at arrhythmia classification with ECGs.

The convergence towards minimal architectural configurations suggests that effective AI-based ECG analysis does not require large computational resources, making deployment in resource-constrained clinical settings feasible. We hypothesise that downsampling and compact representations better capture the relevant features and potentially reveal redundancy in the ECG signal for transformer-based architectures.

### Hyperparameter optimisation

We perform hyperparameter optimisation (input patch size, weighted classes, and loss function) on the NAS output using the PhysioNet 2020 dataset. Variation in patch size (25 to 100 points, corresponding to 0.05 to 0.2 second intervals) has minimal impact on performance, as indicated by minor standard deviations across runs. Classification performance is similar whether model weights are included or excluded from the loss function. This similarity in performance suggests that the data are insufficient for the model to learn class-specific patterns independently.

Regarding loss functions, the BCE loss consistently outperforms the ASL loss. ASL uses a threshold parameter *m* to help models focus on complex cases by down-weighting evident ones. However, whilst *m* can improve focus on challenging examples, it may cause the model to overlook important borderline cases. Using the same threshold across all cardiac arrhythmias assumes equal diagnostic difficulty, which is clinically unrealistic. Without knowledge of the real-world prevalence distributions of arrhythmias, adequately tuning *m* is challenging. A fixed threshold choice likely hinders ASL performance in this context.

Hyperparameter optimisation indicates that class imbalance has the greatest impact on classification performance. None of the hyperparameter choices substantially improved classification performance for rare arrhythmias such as Low QRS Voltage (1.76% of the PhysioNet Challenge 21 dataset) and T-wave inversion (3.24%). These results suggest that class imbalance fundamentally limits classification performance. Data imbalance is particularly problematic for transformer architectures, which lack inductive biases and require extensive training datasets. Convolutional neural networks, for example, allow clinicians and researchers to encode domain knowledge directly into the architecture, such as kernel sizes that reflect the temporal scale of clinically relevant ECG features. Transformer architectures offer no equivalent constraint, learning what to attend to entirely from the data. On an imbalanced dataset, this means the model may preferentially learn features of common classes at the expense of rare ones.

Although the lack of inductive bias makes transformers sensitive to imbalance, the transformer’s self-attention mechanism can learn abstract clinical representations autonomously. This ability is evident since clinical features do not improve classification performance. We hypothesise that the self-attention mechanism may already have learned abstract representations of these features (e.g., heart rate variability from temporal patterns), thereby simplifying clinical deployment by eliminating the need for an explicit feature-extraction pipeline.

## Limitations

The primary limitation of our study is the dataset’s class imbalance. Despite extensive hyperparameter optimisation, we observe no significant improvement in classification performance, with specific rare cardiac abnormalities, such as Low QRS Voltage (1.76%) and T-wave inversion (3.24%), proving particularly difficult to classify.

Although selected for their computational feasibility, popularity, and compatibility with our transformer-based architecture, both AutoFormer and DARTS were originally developed for image classification tasks. As a result, representing the one-dimensional ECG as a two-dimensional image to conform to the AutoFormer search space may not optimally preserve the signal’s temporal characteristics.

## Conclusion

Our NAS revealed that minimal architectural configurations outperform larger alternatives for ECG-based arrhythmia classification. Both macro and micro-level parameters converged toward the smallest values in the search space. The convergence towards minimal architectural configurations suggests that effective AI-based ECG analysis does not require substantial computational resources, making deployment in resource-constrained clinical settings feasible. Given that SCD is sudden and unexpected, prevention strategies in free-living environments require lightweight computational resources suitable for wearable devices. The key insight is that architectural simplicity, rather than complexity, drives classification performance for cardiac abnormalities from ECGs, as adding complexity risks overfitting without improving generalisation.

Hyperparameter optimisation indicates that class imbalance fundamentally limits classification performance, as none of the hyperparameter choices substantially improved classification performance for rare arrhythmias such as Low QRS Voltage and T-wave inversion. Transformer architectures lack inductive biases, making them particularly sensitive to class imbalance. However, the self-attention mechanism can autonomously abstract clinical representations, simplifying clinical deployment by eliminating the need for an explicit feature-extraction pipeline.

Future work could explore custom search spaces tailored to clinical ECG applications, such as incorporating domain-specific signal-processing operations or multi-scale temporal features and investigate how different NAS-derived architectures affect model explainability.

## Data availability

The PhysioNet Challenge 21 is an open-source dataset and can be downloaded from https://physionet.org/content/challenge-2021/1.0.3/#files.

## Contributions

EV: Investigation, visualisation, writing—original draft. AJ: Methodology, software, visualisation, formal analysis. AB, MV: Supervision, writing—review & editing.

